# Real-World Implementation and Clinical Outcomes of Mavacamten in an Underserved Urban Cohort With Obstructive Hypertrophic Cardiomyopathy

**DOI:** 10.64898/2026.09.29.26364363

**Authors:** Joshua Bock, Sandhya Murthy, Aldo L Schenone, Adaya Weissler-Snir

**Affiliations:** Department of Medicine, Jacobi Medical Center, Albert Einstein College of Medicine, Bronx, NY, USA; Division of Cardiology, Montefiore Health System/Albert Einstein College of Medicine, Bronx, NY, USA; Gagnon Cardiovascular Institute, Atlantic Health System, Morristown, New Jersey, USA

**Keywords:** Mavacamten, obstructive hypertrophic cardiomyopathy, Risk Evaluation and Mitigation Strategy (REMS)

## Abstract

**Background:** Real-world data regarding mavacamten use in underserved and racially diverse populations remain limited, particularly for long-term therapy implementation. Structural barriers such as financial instability, fragmented healthcare access, and challenges adhering to REMS-related monitoring requirements may affect longitudinal therapy implementation.

**Methods:** We performed a retrospective observational study of consecutive patients with symptomatic obstructive hypertrophic cardiomyopathy (oHCM) evaluated for mavacamten therapy at a tertiary HCM center in Bronx, NY, between 6/2022-10/2025. Patients were followed for up to 48 weeks after treatment initiation. Clinical characteristics, treatment interruptions, adverse events, echocardiographic and functional outcomes, and implementation barriers were assessed descriptively.

**Results:** Seventy-two patients with symptomatic oHCM were evaluated for mavacamten; sixty-seven initiated treatment. Mean age was 66.9±15.3 years, 72% were female, and 61% identified as Black or Hispanic. Median resting left ventricular outflow tract (LVOT) gradient decreased from 51mmHg (IQR30-88) at baseline to 7mmHg (IQR5-10) at 48 weeks. Median Valsalva LVOT gradient decreased from 74mmHg (IQR50-104) to 20mmHg (IQR10-29). At final follow-up, 80% of patients improved by ≥1 NYHA functional class. LVEF declined below 50% in five patients and recovered in all cases. Approximately one-quarter experienced treatment interruption or discontinuation, nearly half related to insurance instability, non-adherence, and REMS-related monitoring barriers.

**Conclusions:** Mavacamten demonstrated efficacy and safety outcomes comparable to those reported in clinical trials and prior registries in this predominantly underserved minority urban cohort. However, structural and healthcare-access barriers substantially affected long-term treatment implementation. These findings highlight the need to address healthcare-system barriers to ensure equitable implementation of contemporary HCM therapies.

## Introduction

Mavacamten, a first-in-class selective cardiac myosin inhibitor, has demonstrated significant reductions in left ventricular outflow tract (LVOT) obstruction and symptomatic improvement in patients with symptomatic obstructive hypertrophic cardiomyopathy (oHCM) in two pivotal randomized controlled trials.^1,2^ Long-term extension data and the 22-month Risk Evaluation and Mitigation Strategy (REMS) program report have further confirmed its sustained efficacy and favorable safety profile.^3,4^ The 2024 AHA/ACC guideline for the management of HCM recommends mavacamten as Class 1 therapy for patients with oHCM who remain symptomatic (NYHA class 2 or 3) despite standard therapy with beta-blockers or calcium channel blockers.^5^ Despite these advances, data describing the real-world implementation of mavacamten in underserved, racially diverse populations remain scarce. Because these populations have been underrepresented in pivotal clinical trials, understanding the effectiveness, safety, and practical implementation of mavacamten in routine clinical practice is important for ensuring equitable access to this therapy. Barriers to therapy in these communities may include financial instability, language barriers, limited health literacy, inconsistent healthcare access, and difficulty adhering to the monitoring requirements of the U.S. Food and Drug Administration’s Risk Evaluation and Mitigation Strategy (REMS), a drug-safety program designed to ensure that certain medications with serious safety concerns are used safely.^6,7^ These challenges may affect treatment initiation, compliance, and long-term follow-up. We therefore evaluated the implementation, tolerability, and clinical outcomes of mavacamten in a socioeconomically disadvantaged urban population treated at a large tertiary HCM referral center in the Bronx, New York.

## Methods

### Study population

This retrospective cohort study included all patients with oHCM and NYHA class II or III symptoms who were initiated on or referred for initiation of mavacamten at the Montefiore Medical Center HCM clinic between June 2022 and October 2025. Follow-up ended at the time of data extraction, medication discontinuation, death, or loss to follow-up, whichever occurred first.

### Statistical analysis

This descriptive study did not involve formal hypothesis testing or power calculations. Categorical variables are presented as frequencies and percentages, and continuous variables as mean ± standard deviation (SD) or median with interquartile range (IQR), as appropriate. Efficacy parameters were summarized at each follow-up interval for all patients with available data at that time point during extraction. Changes from baseline are reported using mean and median differences with corresponding SD and IQR values. The study was approved by the institutional review board.

## Results

### Baseline characteristics (Table 1)

Seventy-two consecutive patients with symptomatic oHCM were evaluated and considered eligible for mavacamten therapy. Of these, 67 initiated treatment and 5 did not. Among the five patients who did not initiate treatment, three (60%) identified as Black and two (40%) as White. Three patients (60%) had Medicare coverage, one (20%) had Medicaid, and one (20%) was uninsured. The uninsured patient was unable to initiate treatment because of lack of insurance coverage, while the remaining four declined therapy. The 67 patients who initiated mavacamten comprised the study cohort. Mean age was 66.9±15.3 years (range, 21–93 years), 72% were female, and mean follow-up duration was 18.3±10.6 months. The cohort included Black (33%), White (29%), Hispanic (27%), Asian (9%), and other or mixed-race (3%) patients.

**Table 1.** Baseline Characteristics of Patients Initiated on Mavacamten.

| **Characteristic** | **Total (N=67)** |
| --- | --- |
| Age, years (mean ± SD) | 66.9 ± 15.3 |
| Female sex, n (%) | 48 (71.6%) |
| BMI, kg/m<sup>2</sup> (median, IQR) | 29 (25–34) |
| **Race/Ethnicity, n (%)** |  |
| Black | 22 (32.8%) |
| Hispanic | 18 (26.9%) |
| White | 19 (28.4%) |
| Asian | 6 (9.0%) |
| Other | 1 (1.5%) |
| **Insurance payer, n (%)** |  |
| Medicare<sup>^</sup> | 45 (67.2%) |
| Medicaid | 9 (34%) |
| Commercial | 13 (19.4%) |
| **Median household income, n (%)<sup>#</sup>** |  |
| <\$75K | 34 (50.7%) |
| \$75K-\$100K | 11 (16.4%) |
| \$100K-\$125K | 6 (9.0%) |
| \$125K-\$150K | 4 (6.0%) |
| \$150K-\$175K | 4 (6.0%) |
| \$175K-\$200K | 4 (6.0%) |
| \$200K+ | 4 (6.0%) |
| **Comorbidities, n (%)** |  |
| Hypertension | 52 (77.6%) |
| Diabetes mellitus | 15 (22.4%) |
| Hyperlipidemia | 21 (31.3%) |
| Coronary artery disease | 11 (16.4%) |
| Stroke | 4 (6.0%) |
| Chronic kidney disease | 8 (11.9%) |
| Atrial fibrillation (paroxysmal) | 7 (10.4%) |
| Obstructive sleep apnea | 5 (7.5%) |
| Chronic obstructive pulmonary disease | 4 (6.0%) |
| Asthma | 8 (11.9%) |
| Obesity | 6 (9.0%) |
| Ever smoker | 8 (11.9%) |
| Implantable cardioverter-defibrillator, n (%) | 13 (19.4%) |
| **Genetic testing, n (%)** |  |
| Pathogenic/Likely pathogenic variant | 19 (28.4%) |
| Variant of uncertain significance (VUS) | 8 (11.9%) |
| Negative | 27 (40.3%) |
| Not done | 13 (19.4%) |
| **Background therapy at initiation, n (%)** |  |
| Beta-blocker | 46 (68.7%) |
| Calcium channel blocker | 22 (32.8%) |
| Disopyramide | 1 (1.5%) |
| **Echocardiographic parameters** |  |
| Ejection fraction, % (median, IQR) | 70 (65–75) |
| Resting LVOT gradient, mmHg (median, IQR) | 51 (30–88) |
| Valsalva LVOT gradient, mmHg (median, IQR) | 74 (50–104) |
| Left atrium volume index, mL/m<sup>2</sup> (median, IQR)<sup>1</sup> | 45 (40–53) |
| B-type natriuretic peptide, pg/mL (median, IQR)<sup>2</sup> | 203 (105–411) |
| Maximal wall thickness on cardiac magnetic resonance, mm (median, IQR)<sup>3</sup> | 19 (17–20) |
| **NYHA functional class** |  |
| NYHA class II, n (%) | 36 (53.7%) |
| NYHA class III, n (%) | 31 (46.3%) |
| <sup>^</sup> 19 patients had Medicaid as secondary insurance<br><sup>#</sup> American Community Survey 5-year estimates via [unitedstateszipcodes.org](https://www.census.gov/ipeds/data/acs/5year/), per-Zip code data<br><sup>1</sup> n=34 with LAVI; n=62 with BMI recorded.<br><sup>2</sup> n=32 with BNP available.<br><sup>3</sup> n=51 with CMR wall thickness available. |  |

### Treatment discontinuation and interruption (Table 2)

Mavacamten was permanently discontinued in nine patients because of worsening dyspnea, palpitations, or fatigue (n=3), LV systolic dysfunction (n=3), cost (n=2), and loss to follow-up (n=1). Fifty-eight patients remained on therapy, including eight who experienced temporary treatment interruption. Reasons for interruption included reduced LVEF (n=3), non-adherence (n=3), and cost (n=2).

**Table 2.** Non-clinical implementation barriers to mavacamten therapy.

| Barrier | Patients, n | Clinical consequence |
| --- | --- | --- |
| Patient reluctance/refusal | 4 | Therapy not initiated |
| Insurance/cost<sup>a</sup> | 5 | Failure to initiate, treatment interruption, or discontinuation |
| Medication non-adherence | 3 | Temporary treatment interruption |
| Loss to follow-up | 1 | Permanent treatment discontinuation |

<sup>a</sup> Insurance/cost barriers included one patient who did not initiate therapy because of lack of insurance coverage, two who permanently discontinued treatment because of cost, and two who experienced temporary treatment interruption because of cost.

### LVOT gradients (Figure 1)

Median resting LVOT gradient decreased from 51 mmHg (IQR 30–88) at baseline to 18 mmHg (IQR 9–37) at 12 weeks, 14 mmHg (IQR 5–27) at 24 weeks, and 7 mmHg (IQR 5–10) at 48 weeks. The proportion of patients with resting gradients <30 mmHg increased from baseline to 75% at 12 weeks, 84% at 24 weeks, and 96% at 48 weeks. Median Valsalva LVOT gradient decreased from 74 mmHg (IQR 50–104) at baseline to 21 mmHg (IQR 12–49) at 12 weeks, 25 mmHg (IQR 11–52) at 24 weeks, and 20 mmHg (IQR 10–29) at 48 weeks. Correspondingly, the proportion of patients with Valsalva gradients <30 mmHg increased to 65%, 67%, and 76% at 12, 24, and 48 weeks, respectively.

**Figure 1.**
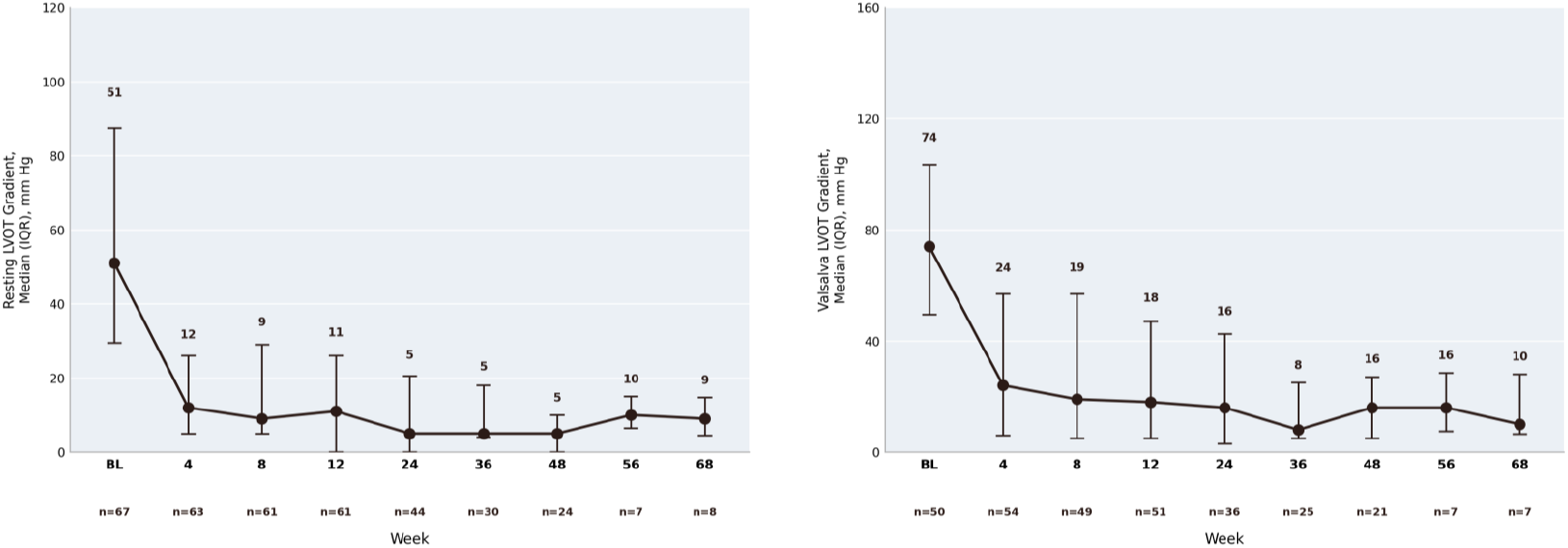
Rest and Valsalva Left ventricular tract outflow gradient over time

### NYHA functional class (Figure 2)

At baseline, 35 patients (52.2%) were in NYHA class II and 32 (47.8%) were in class III. By 48 weeks, patients were equally distributed between class I (50%) and class II (50%), with no patients remaining in class III or IV. At the final available assessment, 80% of patients improved by at least one NYHA class from baseline, including 61.5% who improved by one class and 18.5% who improved by two classes; two patients worsened by one class. The proportion of patients with ≥1 NYHA class improvement was 61% at 12 weeks, 72% at 24 weeks, 81% at 36 weeks, and 73% at 48 weeks.

**Figure 2.**
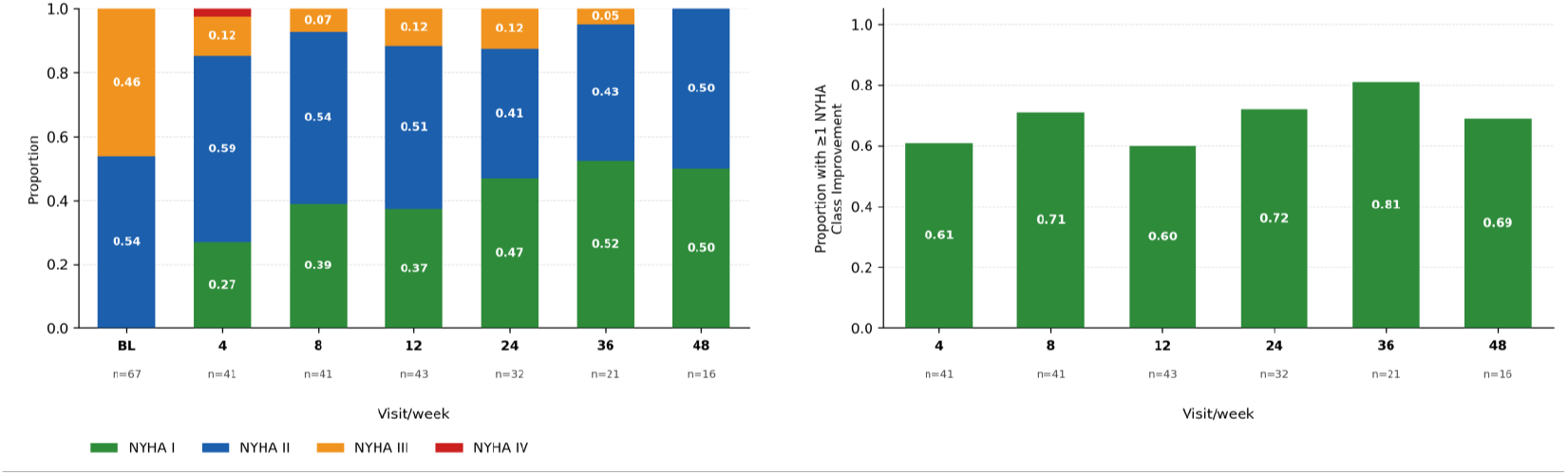
Changes in New York Heart Association (NYHA) class in patients on mavacamten. (A) Proportions of patients at NYHA I, II, III, IV functional classes at Weeks 0, 4, 8, 12, 24, 36 and 48, of mavacamten therapy. (B) Proportion of patients who experienced >1 NYHA functional class improvement compared to their Week 0 NYHA class at Weeks 4, 8, 12, 24, 36 and 48

### Left ventricular ejection fraction (Figure 3)

Median baseline LVEF was 70% (IQR 65–75%). At the final available echocardiogram, median LVEF was 65% (IQR 60–70%), representing a median absolute change of ™5 percentage points (IQR ™10 to 0). LVEF declined below 50% in five patients and recovered in all cases; mavacamten was successfully resumed in two. In two of the five patients, LVEF decline occurred in the setting of concurrent atrial fibrillation (AF) with rapid ventricular response.

**Figure 3.**
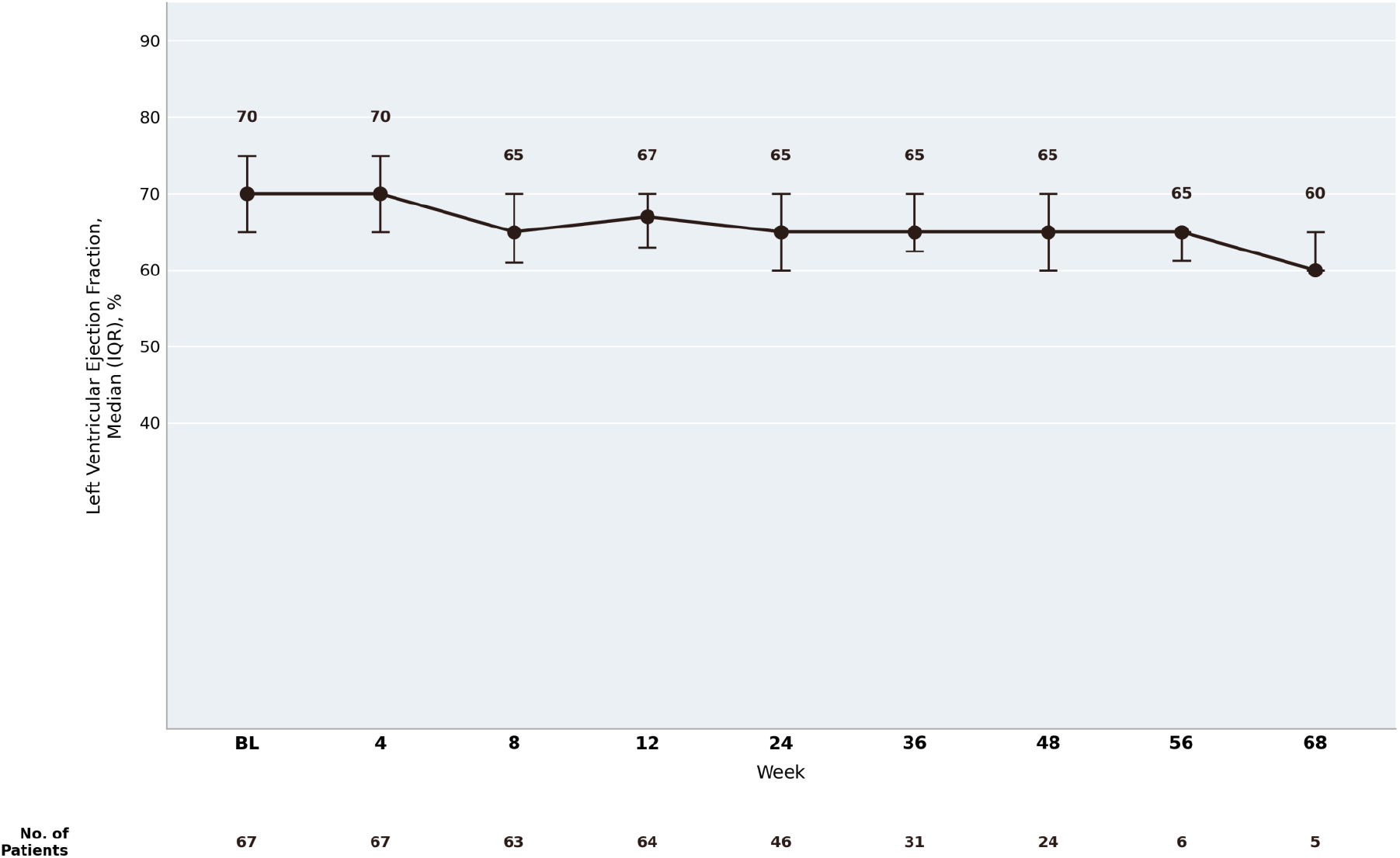
Left ventricular ejection fraction over time

### Adverse events

Adverse events occurred in 22 patients (32.8%). The most common were dizziness or lightheadedness (n=5), palpitations (n=5), and fatigue (n=4). One patient experienced a self-limited episode of supraventricular tachycardia. Worsening dyspnea led to permanent treatment discontinuation in two patients. Among seven patients with baseline paroxysmal AF, four experienced AF with rapid ventricular response, leading to treatment modification in three. One patient progressed to persistent AF and underwent catheter ablation. No new-onset AF occurred in patients without prior AF. There were no mavacamten-related deaths, and no patient had persistent LVEF <50% at final follow-up.

### Background therapy de-escalation

Among the 46 patients receiving beta-blockers at baseline, 17.4% discontinued therapy and 10.9% underwent dose reduction. Among the 22 patients receiving non-dihydropyridine calcium channel blockers, 31.8% discontinued therapy and 9.1% underwent dose reduction. De-escalation of background medical therapy was not associated with recurrent LVOT obstruction or the need for mavacamten dose escalation.

### Drug interactions and medication adjustments

Two patients required substitution of a proton pump inhibitor with an H2 receptor antagonist because of CYP3A4 interactions. One patient receiving tacrolimus required reduced-dose mavacamten initiation with close therapeutic drug monitoring. In another patient receiving venetoclax, mavacamten initiation was deferred until completion of therapy.

## Discussion

This registry extends the growing body of real-world evidence by describing mavacamten use in a predominantly minority, socioeconomically disadvantaged urban population. Unlike prior clinical trials, which predominantly enrolled White patients, more than 60% of our cohort identified as Black or Hispanic.^1,2^ With an age range of 21 to 93 years, including nearly 20% of patients aged ≥80 years, our cohort reflects the breadth of patients encountered in routine HCM practice. Despite substantial demographic and socioeconomic differences from prior studies, the degree of LVOT gradient reduction and symptomatic improvement was comparable to that observed in the pivotal clinical trials^1,2^ and subsequent real-world registries.^3,4,8^ Hemodynamic and symptomatic responses were also similar across age groups. These findings suggest that the efficacy and safety of mavacamten in this diverse real-world cohort were comparable to those reported in pivotal clinical trials and prior registries. However, our experience also demonstrates that successful implementation of mavacamten in underserved populations is influenced as much by social and healthcare-system barriers as by the pharmacologic effects of the drug itself.

### Implementation challenges

Although all patients had been referred to a dedicated HCM center and met clinical criteria for therapy, a proportion declined mavacamten initiation. Among the five patients who did not initiate therapy, three (60%) identified as Black and two (40%) as White; three had Medicare coverage, one had Medicaid, and one was uninsured. Although no clear demographic clustering was apparent, the small number of patients precludes meaningful comparisons. Notably, the uninsured patient was unable to initiate treatment because of lack of coverage, illustrating how financial barriers may prevent access to therapy even before treatment begins. Reluctance to initiate novel therapies requiring intensive monitoring has previously been described in underserved populations and may reflect a combination of medical mistrust, limited representation in clinical trials, concerns regarding medication safety, and the perceived burden of intensive monitoring.^6^ These concerns may be amplified by the requirements of the REMS program, including serial echocardiography and frequent follow-up visits. Clear communication regarding the rationale for monitoring, the reversibility of treatment-related reductions in LVEF, and protocol-driven dose adjustments may improve treatment acceptance. Sustained therapy also proved challenging. Among the 67 patients who initiated mavacamten, 9 (13.4%) permanently discontinued treatment and 8 (11.9%) experienced temporary treatment interruption, yielding a combined rate of 25.4%. This was higher than reported in the pivotal clinical trials and long-term extension studies. In EXPLORER-HCM, temporary interruption and permanent discontinuation due to adverse events occurred in 8.1% and 1.6% of mavacamten-treated patients, respectively.^1^ In the longer-term MAVA-LTE study, 8.7% of patients experienced temporary interruption due to LVEF reduction below 50%, while 5.6% permanently discontinued treatment because of treatment-emergent adverse events over a median follow-up of 166 weeks.^3^ Importantly, eight of the 17 interruptions or discontinuations (47.1%) in our cohort were attributable to non-clinical factors, including cost or insurance barriers (n=4), medication non-adherence (n=3), and loss to follow-up (n=1), representing 11.9% of all treated patients. Although differences in follow-up duration and event definitions limit direct comparisons, these findings underscore that treatment persistence in underserved populations may be substantially affected by healthcare-access and logistical barriers, independently of pharmacologic efficacy or tolerability. Financial barriers persisted despite manufacturer assistance programs, particularly during insurance transitions, and treatment interruptions occurred disproportionately among non-White patients, consistent with broader disparities in healthcare access.^6^ Limited English proficiency and low health literacy further complicated long-term adherence. Although REMS educational materials are available in multiple languages, clinic visits and pharmacy-related communication frequently relied on interpreters, potentially limiting patient understanding of medication monitoring, drug interactions, and the importance of scheduled echocardiographic surveillance. Together, these findings suggest that structural barriers may be as important as clinical factors in determining successful long-term implementation of mavacamten therapy, highlighting the importance of multidisciplinary care, patient education, and longitudinal care coordination.

### Atrial fibrillation

The relationship between cardiac myosin inhibitors and atrial fibrillation (AF) remains uncertain. Pooled analyses of randomized trials have not demonstrated an increased risk of AF compared with placebo, although these studies largely excluded patients with uncontrolled, persistent, or permanent AF.^9,10^ In contrast, a single-center real-world observational study reported a threefold higher risk of new-onset AF than predicted by the HCM-AF score (HR 3.0; P=.01).^11^ Long-term follow-up from MAVA-LTE and data from ODYSSEY-HCM suggest that AF may be more closely associated with transient reductions in LVEF than with an increased incidence of arrhythmia itself.^12,13^ Consistent with these observations, the FDA recommends temporarily interrupting mavacamten in patients who develop uncontrolled AF or other tachyarrhythmias because of the increased risk of systolic dysfunction.^15^

In our cohort, all AF-related events occurred in patients with pre-existing paroxysmal AF, and no patient without prior AF developed new-onset AF during a mean follow-up of 18.3 months. More than half of patients with baseline paroxysmal AF experienced AF with rapid ventricular response, resulting in temporary treatment interruption or dose modification until rhythm or rate control was achieved. In two patients, AF with rapid ventricular response coincided with transient reductions in LVEF below 50%, consistent with observations from MAVA-LTE and ODYSSEY-HCM.^12,13^ Following restoration of rate or rhythm control, LVEF recovered and mavacamten was successfully resumed in all cases. One patient progressed to persistent AF and subsequently underwent catheter ablation, consistent with the natural history of AF in HCM.^14^ Together, these findings suggest that mavacamten did not appear to increase the risk of incident AF in our cohort but that patients with established AF remain susceptible to episodes of rapid ventricular response associated with transient systolic dysfunction, supporting close rhythm surveillance and temporary treatment interruption during uncontrolled AF.

### De-escalation of background therapy

As LVOT obstruction improved, the need for high-dose beta-blockers and calcium channel blockers diminished, with a substantial proportion of patients undergoing dose reduction or complete discontinuation during follow-up, consistent with prior real-world reports.^3-5^ Importantly, these reductions were sustained without subsequent mavacamten dose escalation or recurrence of significant LVOT obstruction, suggesting durable relief of obstruction despite de-escalation of background therapy. These findings highlight an important practical benefit of mavacamten: sustained relief of LVOT obstruction may permit long-term simplification of background medical therapy without loss of hemodynamic benefit, thereby reducing treatment burden and improving medication tolerability in appropriately selected patients.

## Limitations

This study has several limitations. It was a retrospective, single-center observational registry without a control group, limiting causal inference. Missing follow-up data at later time points resulted in smaller sample sizes beyond 48 weeks and may have introduced attrition bias. As the experience of a tertiary HCM referral center, these findings may not be generalizable to all practice settings. Finally, the modest sample size limited subgroup analyses and the ability to detect uncommon adverse events.

## Conclusions

Mavacamten demonstrated efficacy and safety outcomes comparable to those reported in pivotal clinical trials and prior real-world registries in this predominantly underserved, racially diverse urban cohort. However, treatment interruption and discontinuation related to non-clinical factors remained common. Structural barriers and challenges adhering to REMS-related monitoring requirements substantially affected longitudinal therapy implementation. Our findings underscore that addressing healthcare-system barriers may be as important as optimizing pharmacologic therapy to ensure equitable access to contemporary HCM treatment.

## Data Availability

Data will be available upon request

## Notes

### Competing Interest Statement

The authors have declared no competing interest.

### Author Declarations

The study was approved by the institutional review board of Montefiore/Einstein Medical Cetner

